# National assessment of brazilian Community Pharmacists’ knowledge in Medication Dispensing: a cross-sectional survey

**DOI:** 10.1101/2025.06.07.25329194

**Authors:** João Paulo Alves Cunha, João Paulo Vilela Rodrigues, Kérilin Stancini Santos Rocha, Ana Maria Rosa Freato Gonçalves, Fabiana Rossi Varallo, Leoanardo Régis Leira Pereira

**Affiliations:** Pharmaceutical Assistance and Clinical Pharmacy Research Center, Department of Pharmaceutical Sciences, School of Pharmaceutical Sciences of Ribeirão Preto, University of São Paulo (USP), Ribeirão Preto, Brazil; Laboratory for Innovation in Health Care, Graduate Program in Pharmaceutical Sciences, Federal University of Espírito Santo, Vitória, Brazil

## Abstract

Brazilian community pharmacists’ knowledge of medication dispensing was assessed through a nationally representative cross-sectional survey conducted between October 2021 and May 2022. A total of 366 licensed pharmacists working in private community pharmacies—71.3% of whom were female, with an average age of 36.4 years (standard deviation 9.4)—completed a validated online questionnaire covering fundamental aspects of dispensing practice and reported on their undergraduate training and preferred information sources. The overall mean correct-response rate was 70.8%, with the Southeast and South regions scoring highest at 71.0% and 71.9%, respectively. Domains related to antimicrobial and over-the-counter medication dispensing exhibited the largest knowledge gaps across all regions. Graduates of public universities and pharmacists holding postgraduate qualifications demonstrated significantly greater proficiency, and higher knowledge scores correlated with more positive perceptions of the relevance of their training for patient counseling and community health promotion. Internet resources and medication package inserts were the most frequently consulted references for resolving dispensing questions, with 38% of respondents consulting them on a daily basis. These findings reveal a moderate overall level of dispensing competence, pinpoint critical areas for improvement, and underscore the influence of educational background on professional performance. Targeted continuing-education initiatives are therefore essential to reinforce pharmacists’ competencies, enhance the quality of services in community pharmacies, and ultimately improve patient care outcomes.

## Introduction

Community pharmacies are the most available healthcare facilities for the population and are, therefore, strategic locations for developing actions aimed at disease prevention, and health and rational use of drugs promotion. In both public and private community pharmacies, the pharmacist’s daily routine involves technical and managerial activities designed to ensure that medicines, vaccines, and other health products are available to people, in addition to clinical activities. Selecting which medicines to provide based on epidemiological criteria and managing the drug inventory are examples of managerial practices [1,2].

In the context of clinical services within Pharmaceutical Care, health screening, medication reconciliation, pharmacotherapeutic follow-up through pharmaceutical consultations, pharmacotherapy review, monitoring of clinical parameters, and medication dispensing are among the activities that can be carried out in a community pharmacy [2–4]. Drug dispensing can be defined as a clinical service that ensures the provision of drugs through the analysis of technical and legal aspects of prescription, and performance of interventions in the process of medicine use that includes pharmaceutical counseling and documentation of the interventions [2,5].

In the US, it is estimated that 57% of pharmacists work in establishments that perform dispensing [6]. In Brazil, the Federal Pharmacy Council, that regulates the pharmacy profession, indicates that approximately 70% of licensed pharmacists work in community pharmacies. Among them, about 85% practice in private establishments [7].

According to the European Community Pharmacists, the population aging observed in recent decades, and the increase in the prevalence of chronic diseases pose new challenges to health systems worldwide. In this context, the role of pharmacists in activities such as medication dispensing in community pharmacies has great potential to promote adherence to drug therapies, improve health outcomes and people’s quality of life, and reduce the demand for care in higher-complexity health facilities and overall healthcare costs [1]. A systematic review conducted by Pizetta et al., 2021, showed that most of the included studies demonstrated a positive impact of dispensing on the clinical outcomes of different health conditions [8].

The quality of any pharmaceutical service provided is related to the quality of the professional’s education. It is recommended that education for clinical practice be based on the development of competencies that encompass knowledge, skills, and attitudes through practical activities in real-life settings or real-life situations simulation [9,10]. Studies evaluating the knowledge of community pharmacists regarding medication dispensing are scarce, especially in developing countries. Such studies allow for a situational diagnosis and the planning of educational actions to address potential gaps. In this context, the study aimed to assess the knowledge of Brazilian pharmacists working in community pharmacies about medication dispensing, as well as to identify their perceptions of their academic training.

## Methods

### Study Design and Setting

This descriptive, cross-sectional survey was conducted across all Brazilian Federal Units between October 2021 and May 2022. The study followed the recommendations of Kelley et al. (2003) and Bennett et al. (2011) and was reported according to the Consensus-Based Checklist for Reporting of Survey Studies (CROSS) [11, 12].

### Study Participants

All licensed pharmacists with active registration in their respective Regional Pharmacy Councils, working in private community pharmacies in any Brazilian Federal Unit and performing medication dispensing as part of their routine, were invited to participate. No exclusion criteria were applied.

### Sample Size Calculation

According to the Federal Pharmacy Council, Brazil had 234,301 registered pharmacists in 2020. A sample size of 337 pharmacists was calculated for a 5% margin of error and 95% confidence level, considering the finite population. Snowball sampling and convenience selection were used to reach the required sample.

### Data Collection Instruments

The study was carried out by sending two previously validated structured questionnaires online.

#### 1. Questionnaire for the Evaluation of Knowledge about Medication Dispensing (CDM-51) [15]

This questionnaire was developed and validated for Portuguese by Gonçalves and collaborators (2018) and consists of two independent sections. The first refers to the sociodemographic data and schooling of the research participants and has 10 items, while the second part has 51 objective questions, which can be answered with “Yes”, ‘No’ or “I don’t know” and aims to assess the participants’ knowledge about dispensing medicines in six dimensions: 1) Attitudes allowed in the pharmaceutical environment (10 items); 2) Dispensing medicines subject to special control (13 items); 3) Dispensing generic medicines (four items); 4) Dispensing antimicrobials (seven items); 5) Dispensing non-prescription medicines (four items); 6) Dispensing medicines belonging to the Farmácia Popular do Brasil program (government program for access to affordable medicines) and/or used (13 items).

#### 2. Instrument developed by Redigolo (2018) [16]

This instrument assesses the appreciation of training in the Pharmacy undergraduate course, and aims to identify the survey participants’ perception of the usefulness of their training during graduation in carrying out certain professional activities, including dispensing medicines. The instrument is made up of 20 items and uses a six-point Likert Scale to collect responses, with 0 being the lowest point on the scale representing that the participant had no training during graduation for the activity specified in the statement, and 5 being the highest point on the scale representing that the training received by the participant was extremely useful for carrying out the given activity.

For this study, 10 items from this instrument were used. The topics selected were: Filling in the Notification of Irregularity of Medicines suspected of quality deviation or adverse reaction; Delivering medicines; Advising on the availability of items; Informing on the validity of the prescription; Clarifying doubts and/or any other need; Clarifying doubts related to the use of medicines; Guidance on the use of gynecological products; Glycemic monitoring; Pharmacotherapeutic follow-up; General health promotion in the community.

### Research Protocol

Initially, the research instruments were made available in electronic format via Google Forms® and submitted to a pilot study. At this stage, 20 pharmacists with previous experience in dispensing medicines at some point in their professional career took part. The participants answered the instruments in full and, at the end, were able to register their opinions and suggestions about the electronic form, addressing aspects such as layout, average response time, any grammatical errors and the clarity of the questions. Based on the feedback received, minor changes were made, without altering the construct of the instruments, and an average response time of 20 minutes was established.

After this stage, the instruments were disseminated remotely, still using Google Forms®, between October 2021 and May 2022, with the aim of reaching as many pharmacists working in private community pharmacies as possible. The survey was disseminated mainly through social networks (Facebook, Instagram and WhatsApp). Access to the questionnaire was made available both via a QR Code, inserted into graphic materials developed by the researcher to facilitate dissemination, and via a URL (Uniform Resource Locator) link incorporated into the text of posts and shared messages.

In addition, all the Regional Pharmacy Councils in Brazil, as well as the Federal Pharmacy Council, were contacted by telephone and/or e-mail to ask for support in disseminating the survey to pharmacists with active registration.

It should be noted that the questionnaire was anonymous and self-administered, with all the mandatory questions, guaranteeing complete answers. Furthermore, no financial incentives were offered for taking part in the study.

### Statistical Analysis

Continuous variables were presented using mean, standard deviation, minimum and maximum values, while categorical variables were presented using frequency. The Statical Package for Social Sciences® (SPSS) Inc., version 21, 2012 program was used to carry out the statistical analyses. The Poisson model (for comparing the means of discrete variables), Fisher’s exact test (for assessing the association between categorical variables) and the ANOVA test (for comparing the score and the degree of usefulness of the degree between the regions) were used to evaluate the variables between the regions. The comparison of pharmacists’ knowledge between sociodemographic variables and perception of academic training was carried out using Student’s t-test and ANOVA test (for categorical independent variables) and Pearson’s correlation coefficient (for quantitative independent variables). A significance level of 5% was set. The results were considered statistically significant when p ≤ 0.05.

### Ethical Aspects

The Research Ethics Committee of the Faculty of Pharmaceutical Sciences of Ribeirão Preto (FCFRP-USP) approved the study (CAAE: 34271520.3.0000.5403; Opinion No. 5.058.567).

Informed consent was obtained electronically prior to participants accessing the questionnaire. Participants were presented with an online consent form detailing the study’s objectives, procedures, potential risks, and confidentiality measures. Only those who actively selected the ‘I agree to participate’ option were granted access to the questionnaire. All participants were 18 years of age or older. No minors were involved in the study.

## Results

### Sample Characterization

A total of 366 pharmacists were included, representing all Brazilian regions, with the majority from the Southeast (52.5%; n = 192) and Northeast (24.6%; n = 90). The sample was predominantly female (71.3%; n = 261), a trend observed across all regions. The national mean age was 36.4 ± 9.4 years (range: 22–70).

Table 1 presents participants’ academic backgrounds. The variables “Educational Qualification” (p = 0.01), “Years Since Pharmacy Graduation” (p < 0.001), and “Years of Professional Experience” (p < 0.001) differed significantly by region. Pharmacists in the South had the longest time since graduation, while those in the South and Southeast also reported the greatest dispensing experience.

**Table 1.**
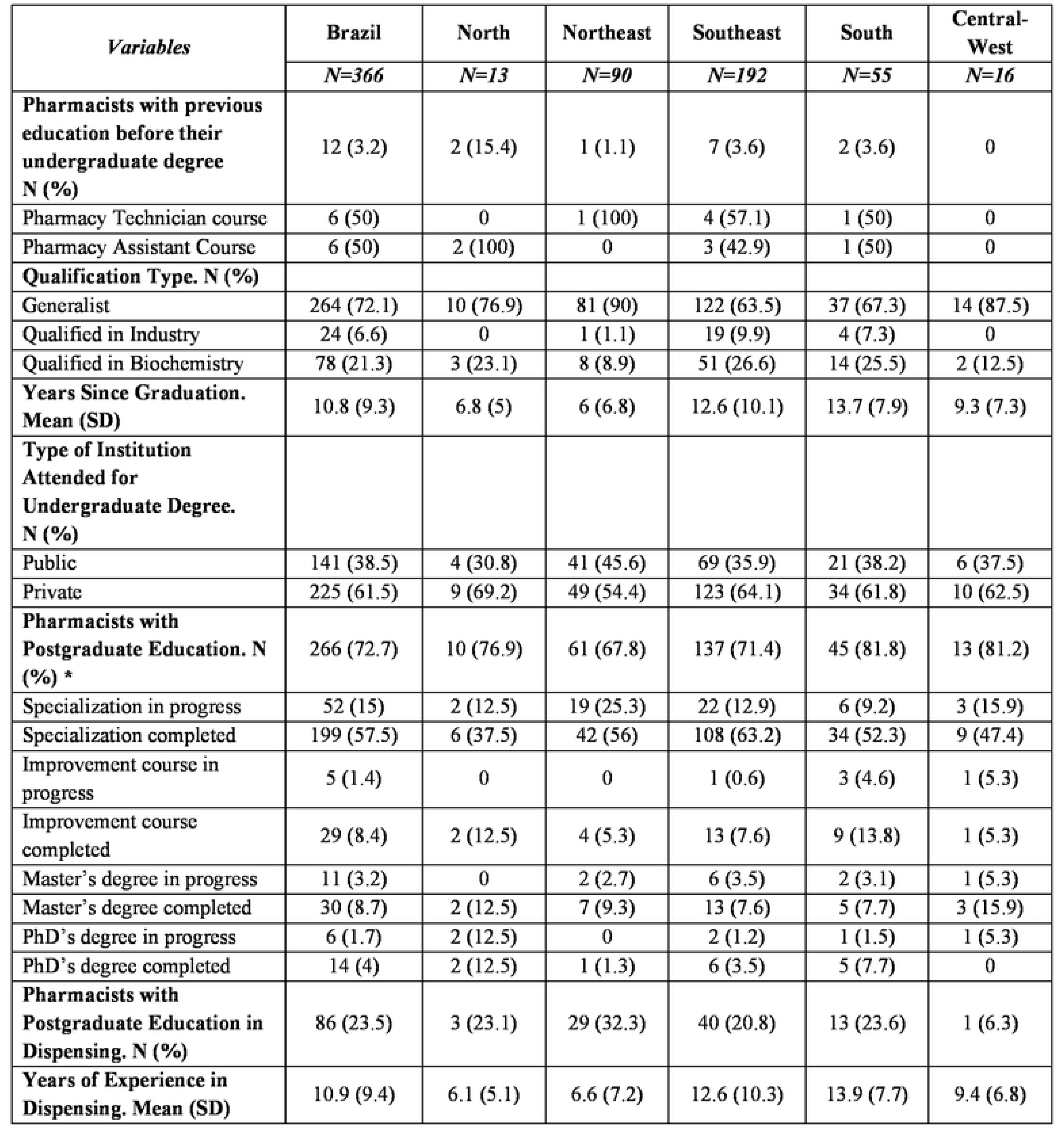
Educational profile of the pharmacists who participated in the study.

Most pharmacists (97%; n = 355) reported having encountered questions about medications, with 100% of participants in the North confirming this. The most frequently used resources for clarification were the Internet (44.2%) and drug package inserts (21%), showing similar patterns across regions. Among those reporting doubts, 38% (n = 135) consulted these resources daily.

### Knowledge Assessment

The national mean percentage of correct responses on the CDM-51 was 70.8%. National and regional means are depicted in Figure 1. No significant differences were observed between regions (p = 0.868).

**Figure 1.**
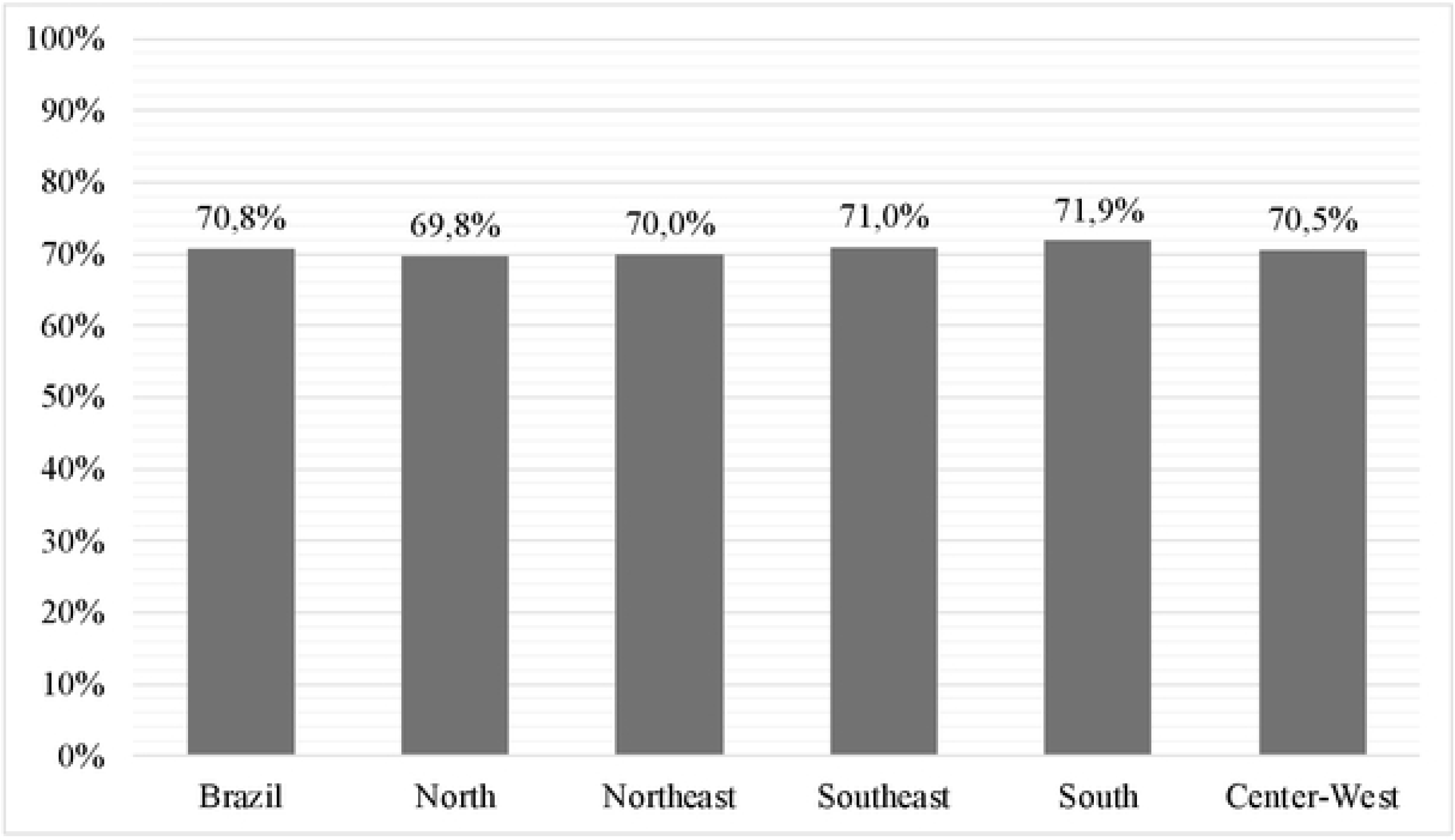
Regional and National Averages of the Percentage of Correct Answers on the CDM-51.

When examining each CDM-51 dimension separately, all showed mean scores above 50%. Two regions exceeded the national overall score: the Southeast (71%) and the South (71.9%), the latter achieving the highest regional score. Table 2 details pharmacists’ percent-correct scores by region across the CDM-51 dimensions. No significant interaction was found between dimensions and region (p = 0.751).

**Table 2.** Regional and national averages of the percentage of correct answers in the domai ns comprising the COM-51.

| *Domains (CDM-51)* | **Brazil** | **North** | **Northeast** | **Southeast** | **South** | **Central-West** |
| --- | --- | --- | --- | --- | --- | --- |
|  | *N=366* | *N=13* | *N=90* | *N=192* | *N=55* | *N=16* |
| Permitted Attitudes in the Pharmaceutical Environment (D1) | 68.9% | 73.1% | 67.8% | 69.3% | 68.0% | 68.8% |
| Dispensing of Controlled Substances (D2) | 72.1% | 74.0% | 69.1% | 72.5% | 76.1% | 69.7% |
| Dispensing of Generic Medicines (D3) | 78.0% | 75.0% | 76.7% | 78.4% | 78.6% | 81.3% |
| Dispensing of Antimicrobials (D4) | 67.0% | 63.7% | 69.8% | 66.6% | 65.5% | 63.4% |
| Dispensing of Over-the-Counter Medicines (D5) | 64.7% | 63.5% | 66.1% | 63.9% | 54.2% | 56.3% |
| Dispensing of Medicines from the Brazilian Popular Pharmacy Program and/or Commonly Used Medicines (D6) | 72.8% | 66.9% | 71.7% | 73.2% | 73.4% | 77.4% |

Correlations between CDM-51 scores and other study variables are presented in Table 3.

**Table 3.** Correlation Between the Average Score on the CDM-5I and Variables Related to the Training of Community Pharmacists.

| *Variables* | **Brazil**<br>**(N=366)** | **Mean Score**<br>**CDM-51** | *p* |
| --- | --- | --- | --- |
| **Pharmacists with a course prior to their undergraduate degree. N (%)** |  |  |  |
| No | 354 (96.8) | 36.14 | 0.510 |
| Yes. Pharmacy Technician Course | 6 (1.6) | 33.83 |  |
| Yes. Pharmacy Assistant Course | 6 (1.6) | 36.12 |  |
| **Type of Qualification. N (%)** |  |  |  |
| Generalist | 264 (72.1) | 36.11 | 0.509 |
| Qualified in Industry | 24 (6.6) | 37.33 |  |
| Qualified in Biochemistry | 78 (21.3) | 35.81 |  |
| **Type of Institution Attended for Undergraduate Degree. N (%)** |  |  |  |
| Public | 141 (38.5) | 37.04 | 0.014* |
| Private | 225 (61.5) | 35.55 |  |
| **Pharmacists with Postgraduate Education. N (%)** |  |  |  |
| Yes | 266 (72.7) | 36.98 | 0.001* |
| No | 100 (27.3) | 33.85 |  |
| **Pharmacists with Postgraduate Education in Dispensing. N (%)** |  |  |  |
| Yes | 86 (23.5) | 37.52 | 0.01* |
| No | 280 (76.5) | 35.75 |  |
| * p<0.05 |  |  |  |

### Perception of Academic Training

Descriptive results regarding community pharmacists’ perceptions of their undergraduate training for medication-dispensing practices and related activities are shown in Figure 2.

**Figure 2.**
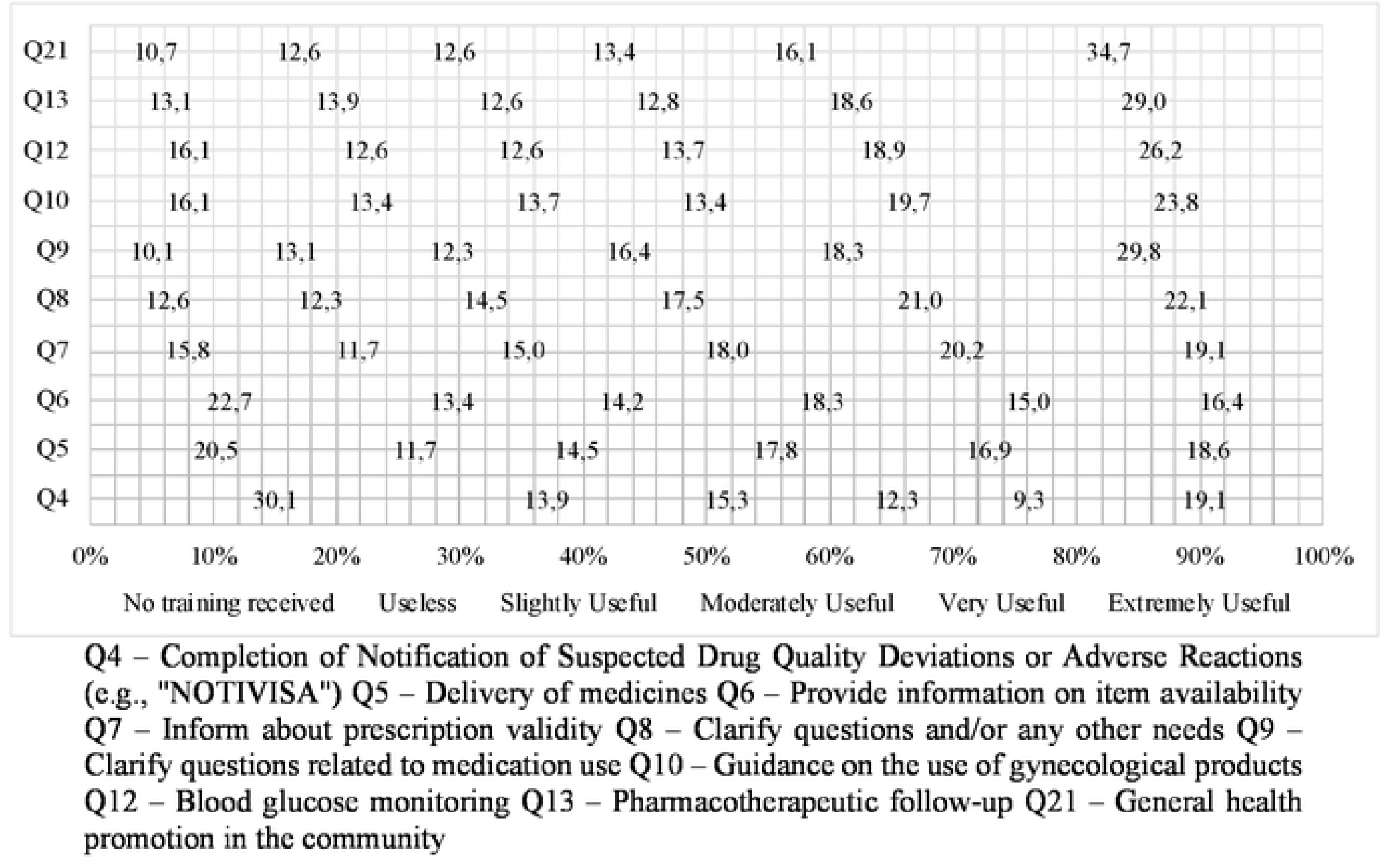
National sample’s perception of the usefulness of the Pharmacy Degree for performing activities related to medication dispensing.

Associations between CDM-51 scores and participants’ evaluations of the impact of their undergraduate education on dispensing activities are summarized in Table 4. Clarifying medication-use questions (Q9) and general health promotion in the community (Q21) were both significantly correlated with CDM-51 scores (p < 0.05). No other significant correlations were found between training perception and the sociodemographic or professional variables investigated (p > 0.05).

**Table 4.** Statistical analysis of possible correlations between the mean score on the CDM-51 and the perception of training for performing activities related to Medication Dispensing.

| *Activities* | *p* |
| --- | --- |
| Completion of Notification of Suspected Drug Quality Deviations or Adverse Reactions (e.g., "NOTIVISA") (Q4) | 0.281 |
| Delivery of medicines (Q5) | 0.511 |
| Guidance on item availability (Q6) | 0.399 |
| Information about prescription validity (Q7) | 0.068 |
| Clarification of questions and/or any other needs (Q8) | 0.058 |
| Clarification of questions related to medication use (Q9) | 0.01* |
| Guidance on the use of gynecological products (Q10) | 0.237 |
| Blood glucose monitoring (Q12) | 0.112 |
| Pharmacotherapeutic follow-up (Q13) | 0.224 |
| General health promotion in the community (Q21) | 0.027* |
| *p<0.005 |  |

## Discussion

According to the Federal Pharmacy Council, approximately 60% of pharmacists with active registration work in commercial pharmacies [17], which underscores the notable relevance of studies evaluating these professionals’ knowledge and practice. The analysis of private community pharmacists’ knowledge regarding the medication-dispensing process at a national level was unprecedented. This research included pharmacists from all Brazilian regions—a critically important consideration in a country of continental dimensions and marked social, cultural, and economic disparities between regions [18].

Given that the CDM-51 comprises items related to content addressed in the 2002 and 2017 National Curriculum Guidelines (DCN), and those Brazilian pharmacists achieved a mean correct-response rate of 70.8% (Figure 1), there is clear need for initiatives to enhance pharmacists’ qualifications in medication dispensing to fill existing knowledge gaps [10, 19]. Between 2019 and 2024, the Brazilian Ministry of Health provided training in medication dispensing and Pharmaceutical Care to pharmacists in public community pharmacies [20]. Our results suggest that similar strategies targeted at private community pharmacists should be considered.

When examining CDM-51 performance by domain, Domain 4 (dispensing of antimicrobials) and Domain 5 (dispensing of over-the-counter medicines, OTC medicines) yielded the poorest results, being the only domains with scores below 70% across all Brazilian regions (Table 2). The World Health Organization estimates that antimicrobial resistance directly caused approximately 1.3 million deaths worldwide in 2019 and contributed to an additional 4.9 million deaths that same year [21]. From a global economic perspective, the World Bank reports that managing health events and outcomes associated with antimicrobial resistance cost up to USD 200 billion in 2022, with projections indicating these costs may reach USD 1.2 trillion by 2050 [20]. Robust evidence shows that inclusion of pharmacists in antimicrobial stewardship teams promotes rational antimicrobial use, improves health outcomes, and reduces costs— particularly in hospital and emergency-care settings [22–25].

Although the opportunity for private community pharmacists to serve on antimicrobial stewardship teams is more limited than in hospitals, appropriately conducted antimicrobial dispensing by clinically trained pharmacists can positively impact antimicrobial-resistance indicators and clinical outcomes, thereby reducing healthcare costs. The potential impact of community pharmacists in promoting rational antimicrobial use has been documented by several authors [26–28].

Regarding OTC medicines, Mota et al. (2020) corroborated our findings by highlighting the clinical and legal risks arising from insufficient knowledge about OTC products [29]. Use of OTC medicines without guidance from a qualified healthcare professional can harm patients, who often underestimate the risks associated with medications that do not require a prescription [30]. Our findings underscore the need for policies that establish educational strategies both for the initial training of pharmacy students and for continuing-education programs, aiming to strengthen community pharmacists’ clinical competencies in health-service provision, including dispensing of antimicrobials and OTC medicines.

In the correlation analysis between CDM-51 mean scores and training-related variables (Table 3), pharmacists holding postgraduate qualifications performed significantly better on the CDM-51 (p < 0.001). Among these, approximately one third (32.3%) reported completing a clinically oriented postgraduate program that included medication-dispensing content. Similar results have been reported in a systematic review emphasizing the importance of educational strategies targeting pharmacists in clinical practice and medication dispensing [31]. Moreover, pharmacists graduating from public universities achieved higher knowledge scores in medication dispensing (p = 0.014). Reis et al. (2015) observed the same profile among participants, suggesting potential gaps in the teaching–learning process at private higher-education institutions [32]. Indeed, Brazil has approximately 72 public pharmacy programs compared to 725 in private institutions, reflecting considerable heterogeneity in the training of future community pharmacists [33].

Data from the Ministry of Education’s Pharmacy Course Report, which includes results from the National Student Performance Exam (ENADE), indicate that public institutions scored higher than private ones [33]. These findings reinforce the need for strategies to improve teaching quality at private pharmacy schools while maintaining high standards in public institutions, for example by incorporating student engagement in practical scenarios and problem-based discussions grounded in active-learning methodologies and competency-based education [10].

It is also important to consider that human and material resources are essential for implementing and sustaining educational strategies. A curriculum with insufficient credit hours, low faculty training, and inadequate practical infrastructure may constitute barriers to high-quality pharmacy education, ultimately reflected in suboptimal professional performance of graduating pharmacists [34,35].

This study further revealed that the sources pharmacists consult when questions arise during dispensing are of low quality, a finding consistent with other Brazilian studies [36–40]. This context highlights the importance of encouraging pharmacists to seek information from robust sources, since pharmacies provide health-related services— including medication dispensing, which has clinical, humanistic, and economic impacts on patient care [8].

Finally, a substantial proportion of pharmacists reported having received little or no training in medication dispensing or that their training was not useful (46.7%; Q5, Figure 2). Better perceptions of training for medication-use counseling and community health promotion were directly associated with higher CDM-51 scores (Table 4). These results confirm the necessity of continuing education in dispensing and suggest the importance of assessing community pharmacists’ needs and knowledge gaps.

This study has limitations that preclude exhaustive analysis of all possible factors, indicating opportunities for future research, such as examination of additional professional variables and classification of pharmacists’ knowledge levels using the CDM-51 instrument.

## Conclusion

The mean correct-response rate in the knowledge assessment of Brazilian pharmacists regarding medication dispensing was 70.8%. Training at public institutions and completion of postgraduate education were associated with higher knowledge levels. Considering that dispensing is one of the primary services provided to the public in community pharmacies, it is essential to implement strategies—including continuing education—to strengthen pharmacists’ qualifications and, consequently, improve the quality and effectiveness of the service delivered.

## Data Availability

All relevant data are within the manuscript and its Supporting Information files.

## Notes

### Competing Interest Statement

The authors have declared no competing interest.

### Funding Statement

The author(s) received no specific funding for this work.

### Author Declarations

The Research Ethics Committee of the Faculty of Pharmaceutical Sciences of Ribeirão Preto (FCFRP-USP) approved the study (CAAE: 34271520.3.0000.5403 Opinion No. 5.058.567).

